# Exploring patient acceptability of emerging intravitreal therapies for Geographic Atrophy: a mixed-methods study

**DOI:** 10.1101/2022.09.19.22279938

**Authors:** Jamie Enoch, Arevik Ghulakhszian, Mandeep Sekhon, David P. Crabb, Deanna J. Taylor, Christiana Dinah

## Abstract

**Purpose:** The acceptability of emerging intravitreal therapies for patients with Geographic Atrophy (GA) is currently unknown. This study therefore aimed to: investigate whether regular intravitreal injections will be acceptable as treatment for GA patients; identify which attributes of current treatments in late stage development patients find less acceptable; and explore whether patient-related factors influence GA treatment acceptability.

**Design:** Exploratory, cross-sectional, mixed-methods study.

**Participants:** 30 UK-based individuals with GA secondary to age-related macular degeneration (AMD), recruited from two London-based hospitals, interviewed in April-October 2021.

**Methods:** Participants responded to a structured questionnaire, as well as open-ended questions in a semi-structured interview. Quantitative data were analysed using descriptive statistics and non-parametric measures of correlation. Qualitative data were analysed using the framework method of analysis, informed by the Theoretical Framework of Acceptability.

**Main outcome measures:** Main quantitative measures were Likert-type scale responses about acceptability of GA treatments. Qualitative outcomes of interest related to participants’ hopes, concerns and understanding of the proposed new intravitreal treatments for GA.

**Results:** Twenty participants (67%) were female, and median (interquartile range (IQR)) age was 83 (78, 87) years. 37% of participants had foveal centre-involving GA, and better eye median (IQR) logMAR visual acuity was 0.30 (0.17, 0.58). Data suggested that 18 participants (60% (95% CI: 41-79%)) would accept the treatment if offered today, despite their awareness of potential drawbacks. Eight participants (27% (95% CI: 10-43%) were ambivalent or undecided about treatment, and four (13%) (95% CI: 0-26%) would be unlikely to accept treatment. Reducing the frequency of injections from monthly to every other month increased the proportion of participants who considered the treatments acceptable.

Qualitative data indicated that participants’ prioritisation of continuation with vision-specific activities influenced treatment acceptability. Conversely, factors limiting acceptability clustered around: the limited magnitude of treatment efficacy; concerns about side effects or the increased risk of neovascular AMD; and the logistical burden of regular clinic visits for intravitreal injections. Misunderstandings of potential benefits indicate the need for appropriately designed patient education tools to support decision-making.

**Conclusions:** Our study suggests a majority of participants would be positive about intravitreal treatment for GA, in spite of potential burdens.

## Introduction

Geographic Atrophy (GA) is the advanced form of the non-neovascular (‘dry’) type of age-related macular degeneration (AMD). In GA, progressive loss of areas of the retinal pigment epithelium, photoreceptors and underlying choriocapillaris occur leading to irreversible vision loss. Estimates suggest there are more people in the United Kingdom living with GA (276,000) than with neovascular (‘wet’) AMD (263,000),^1^ and GA accounts for approximately one-quarter of legal blindness in the UK.^2^ Globally, around 5 million people have GA in at least one eye,^3^ and the incidence is expected to rise with ageing populations. About one-half of patients develop GA in both eyes within seven years of initial diagnosis.^4^ In addition, up to 25% of eyes presenting with wet AMD may have concurrent geographic atrophy at baseline.^5^ While there are now approved treatments for wet AMD, there is currently no therapy for GA, a significant unmet need.

GA is associated with a decline in reading speed.^6^ Even before the foveal centre is involved, GA can have significant impact on functional activities and vision-related quality-of-life,^7^ including difficulties with face recognition,^8^ visual search,^9,10^ and greater anxiety about mobility relative to visually healthy peers.^11^ Furthermore, the condition can impact upon the psychosocial health and wellbeing of the person beyond the effect on vision alone, even before functional vision is significantly affected.^12–15^ Indeed, qualitative studies on the experience of living with GA suggest that many individuals living with the condition feel hopelessness and despair at the prospect of their vision deteriorating, particularly due to the lack of any treatment that could delay, halt or reverse this process.^12^

Dysregulation of the complement cascade has been implicated in the pathogenesis of GA, and there are now two intravitreal complement inhibitors in late-stage development for the treatment of GA.^16^ Regular intravitreal injections are the standard of care for wet AMD, and a common mode of delivery in the current pipeline of treatments for GA in clinical trials. Recent positive results from phase 3 clinical trials of two intravitreal complement inhibitors (Table 1) provide hope for a treatment for GA. However, it is not yet known whether these treatments will be acceptable to patients outside clinical trial settings.

**Table 1.**
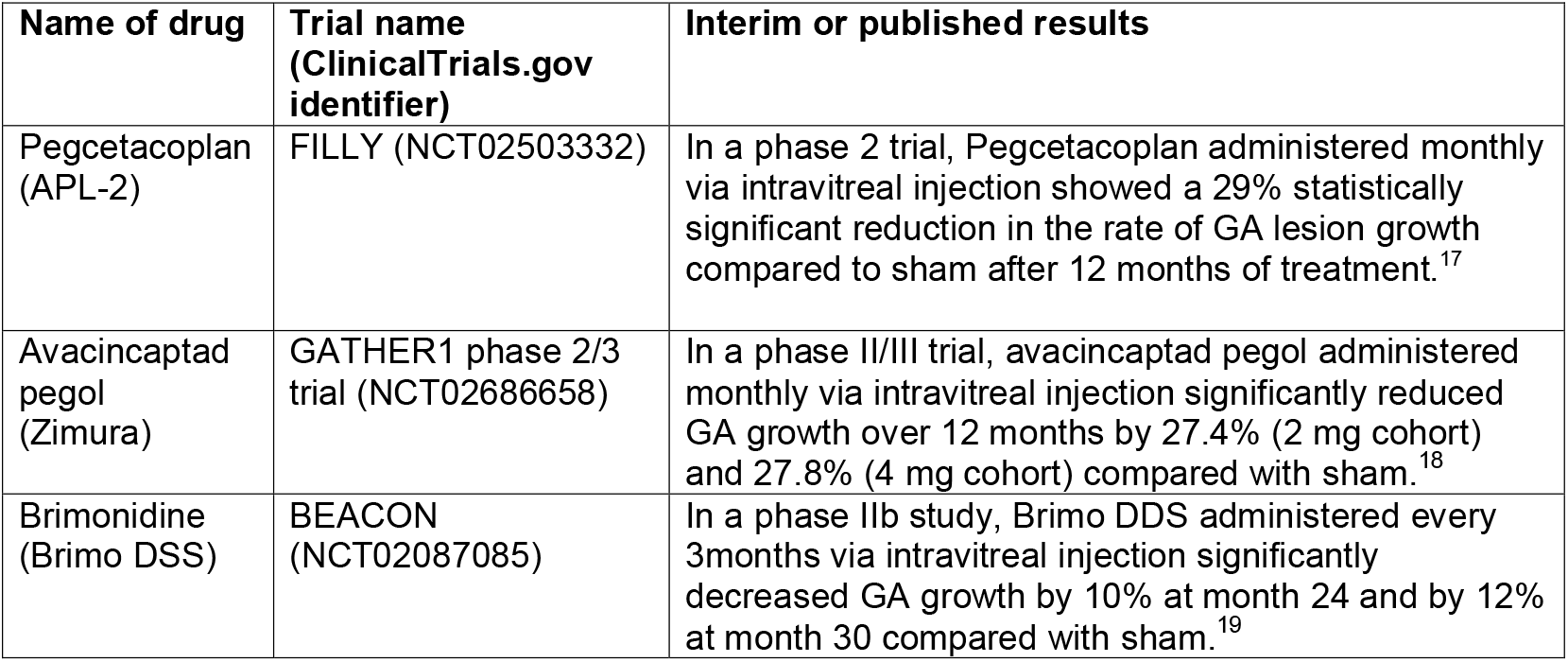
Summary of phase 2 clinical trial results of three investigational intravitreal therapies for GA that were available at the time of study design. ^17 18 19^

Current evidence from wet AMD suggests people will persevere with regular intravitreal treatment, even when associated with a high burden, when motivated by outcome expectations.^20, 21^ Despite efficacious outcomes of anti-VEGF therapy,^22^ some wet AMD patients report significant treatment burden associated with regular intravitreal injections, not only in terms of anxiety, discomfort, pain and/or side effects associated with these injections, but also the logistics of regularly travelling to the eye clinic, waiting times, and impacts on accompanying relatives or caregivers.^23–25^

However, GA is different to wet AMD, being slower to progress, with well-documented variation in rates of progression across individuals, and asymptomatic in some patients until involving the fovea.^26,27^ Therefore, it is vital to understand whether patients with GA would find it acceptable to commence and adhere to frequent intravitreal treatments, in order to slow GA progression. Furthermore, identifying barriers and facilitators that influence the acceptability of such treatments – even prospectively before they are approved - could help to address modifiable factors, and thereby improve treatment delivery and service design.

Acceptability, as defined by Sekhon and colleagues in their Theoretical Framework of Acceptability (TFA), is a “multi-faceted construct that reflects the extent to which people delivering or receiving a healthcare intervention consider it to be appropriate, based on anticipated or experienced cognitive and emotional responses to the intervention”.^28^ Acceptability is a crucial yet complex factor which can have implications for patients deciding to undergo a treatment, as well as adhering and persisting with it. As such, assessment of acceptability to patients should be a critical first step in the design, evaluation and delivery of healthcare interventions.^29^

Our study’s central objective was to explore the overall acceptability of current intravitreal treatments in late-stage development for a sample of GA patients. We aimed to identify which aspects of the treatment are considered less acceptable; and to understand whether specific patient-related factors, contexts and circumstances influence GA treatment acceptability. A secondary aim was to explore what people with GA understand about their disease, its progression, current service provision, and their hopes for GA treatment and/or cure.

## Methods

### Study design and procedure

This study employed a cross-sectional, mixed-methods design,^30^ and full detail on methodological aspects is presented in the published study protocol.^31^ In summary, it was deemed important to quantify participants’ attitudes to acceptability, but also to integrate qualitative data through open-ended questions, in order to explore participants’ perspectives in-depth and understand their beliefs, hopes and concerns regarding GA treatment within their unique contexts and circumstances.

The study procedure is summarised in Figure 1. Before providing informed consent to participate, participants read a leaflet containing details about GA, typical progression and an overview of the current treatments in late-stage development, in terms of mode of delivery, interval of treatment, efficacy outcomes and side effect profiles. After informed consent was obtained, the interviewer (AG, JE or CD) obtained demographic information and administered the EQ-5D, a standardised, validated instrument for patients to self-report their overall health.^32^ A series of Likert-type scale questions regarding knowledge and concerns about GA were administered, before background information about new treatments for GA was reiterated and discussed with participants. Subsequent Likert-type scale questions focussed on the acceptability of new treatments. Next, participants were asked open-ended questions to explore their thoughts on the acceptability of emerging treatments for GA, and the advantages and disadvantages of treatment. The interview schedule, including Likert-type scale questions and semi-structured open-ended questions, is shown in Appendix 1. This interview schedule was developed in consultation with a group of eight patient advisors, individuals living with GA who did not participate in this study but generously volunteered their time and insights.

**Figure 1.**
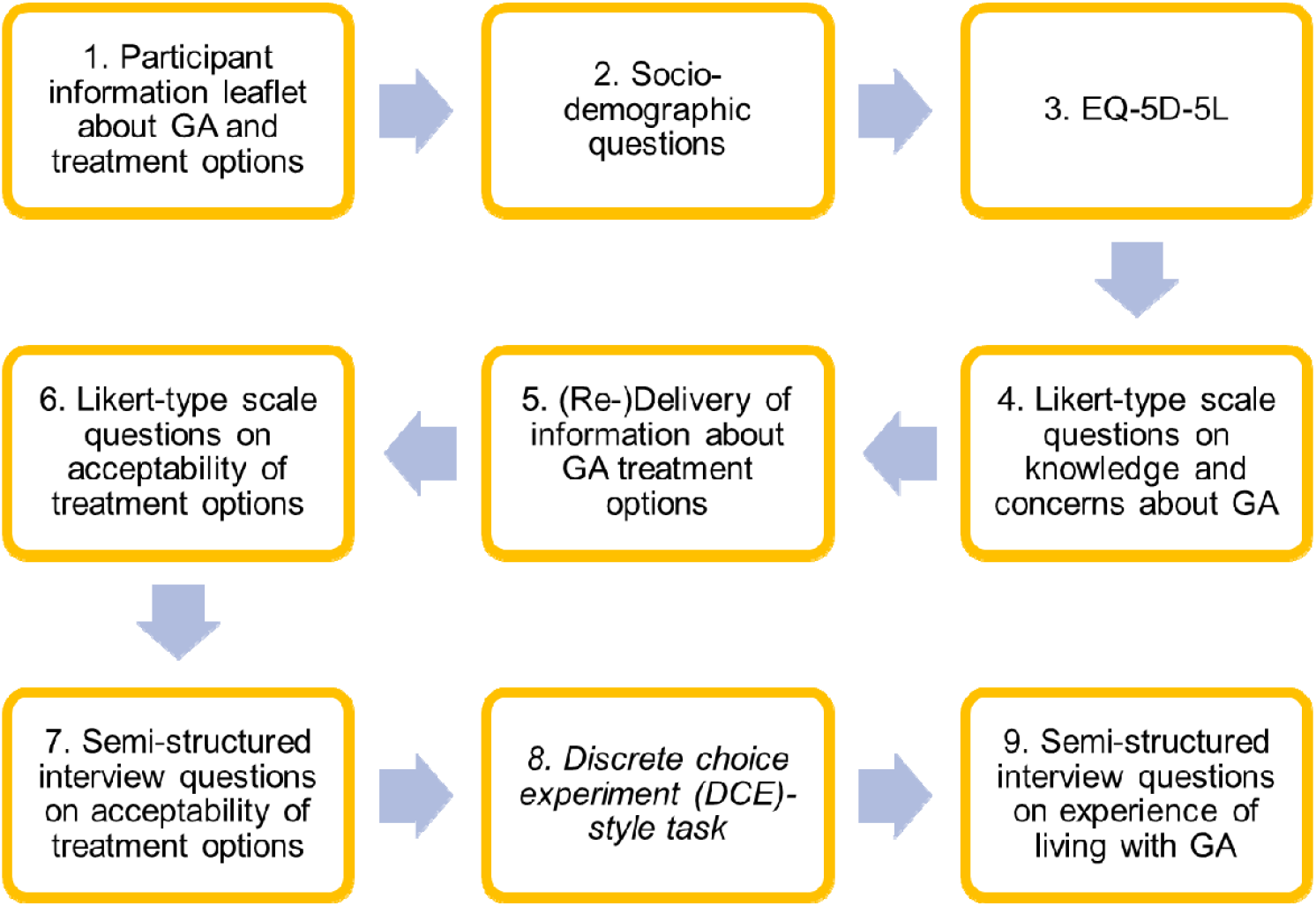
Summary of study procedure

Step 8 (Figure 1) was a task inspired by Discrete Choice Experiments;^33^ this data will be presented in a separate paper.

### Participant recruitment

Individuals with a diagnosis of GA were recruited from two Medical Retina clinics in Brent, one of the most ethnically diverse boroughs in London, UK.^34^ Included participants were required to be aged ≥ 50 years, and have a diagnosis of GA (bilateral or unilateral) secondary to age-related macular degeneration. Patients with other causes of GA - such as Stargardt’s - or with concurrent retinal conditions were excluded. The aim was to recruit a cohort representative of the population in the community; therefore, some participants required an accompanying relative/caregiver to interpret parts of the interview.

In order to explore the views of participants with varied demographic and clinical characteristics, a purposive sampling strategy was employed, aiming to achieve maximum variation^35^ in terms of: age; gender; ethnicity; education level; overall health status; prior experience of intravitreal injections (for wet AMD); best-corrected visual acuity (BCVA); laterality; and foveal involvement, with extrafoveal defined as greater than 0 microns from the fovea.^36^

Consenting participants undertook an audio-recorded interview face-to-face or via telephone with authors AG, CD or JE between April and October 2021. This decision to undertake certain interviews by telephone was a pragmatic response to COVID-19 restrictions in place in the UK at the time.^37^

### Ethical considerations

Ethics Committee approval was obtained from the NHS Health Research Authority on 23 March 2021 (IRAS Project ID: 287824), and the study adhered to the tenets of the Declaration of Helsinki.

### Data analysis

#### Quantitative responses

Descriptive analysis of demographic information and responses to the Likert-type scale questions was undertaken. Where appropriate, Spearman’s rank (*r*_s_) correlation coefficients were calculated to explore potential associations between responses to the Likert-type scale questions on acceptability (dependent variables) and demographic and clinical characteristics (independent variables). A *p-*value of <.05 was considered statistically significant. Statistical tests were conducted using SPSS, version 27.0 (SPSS Inc., Chicago, IL, USA).

#### Qualitative responses

Data from the semi-structured interview were transcribed verbatim, and analysed using the Framework Method of analysis.^38,39^ This systematic qualitative data analysis method allowed for both inductive analysis (whereby open coding of the data leads to generation of themes) and deductive analysis (whereby pre-existing theory – in this case, the TFA - shapes the development of themes). Initial coding was conducted by author JE, followed by a second round of coding involving authors JE, AG, DJT and CD working collaboratively. Discrepancies regarding the best fit of text segments within the TFA matrix were resolved by author MS, an expert in acceptability who developed the TFA. This was an iterative, recursive process, and over time the team collaboratively developed a codebook (Appendix 2), establishing decision rules for coding the data into the seven TFA constructs. The software package NVIVO V.10.2 (QSR International, Cambridge, Massachusetts, USA) was used to manage the qualitative data.

In tandem, data which did not fit within a TFA construct were coded inductively by authors JE, AG and CD, to develop a second framework matrix encapsulating important patterns in the data falling outside the TFA.

Analysis of qualitative data within the framework matrix illustrated that participants’ responses fell within three distinct and recognisable positive, ambivalent, and negative categories.^40^ The categorisation was based on participants’ expressed intentions regarding the potential treatments. For example, a participant concluding that “*I think I would have the treatment at almost any cost*” (P26) would be placed in the positive category, while a participant concluding that the treatment “*is not for me*” (P24) would be placed in the negative category. Two authors (CD and JE) independently assigned the participants into the three categories, and then compared and collaboratively refined the categorisation. Certain disagreements in categorisation were discussed with reference to the individual case in the framework matrix, and all authors subsequently met to consider these disputed cases and reach consensus. After whole team discussion, the three categories were termed “Treatment at any cost” (positive), “Ambivalent”, and “Unlikely to Proceed” (negative).

## Results

### Participants

Thirty participants (67% female) were interviewed, and demographic and clinical characteristics for each participant are displayed in Appendix 3. Median (interquartile range (IQR)) age was 83 (78, 87) years. Nineteen (63%) of participants identified as white, eight (27%) as South Asian, one (3%) as Black, and two (7%) as another ethnicity. The range of participants’ primary languages is displayed in Figure 2. In the case of three participants (P16, P20, and P25), interviews were interpreted by or mediated through an accompanying relative, due to English language or communication difficulties.

**Figure 2.**
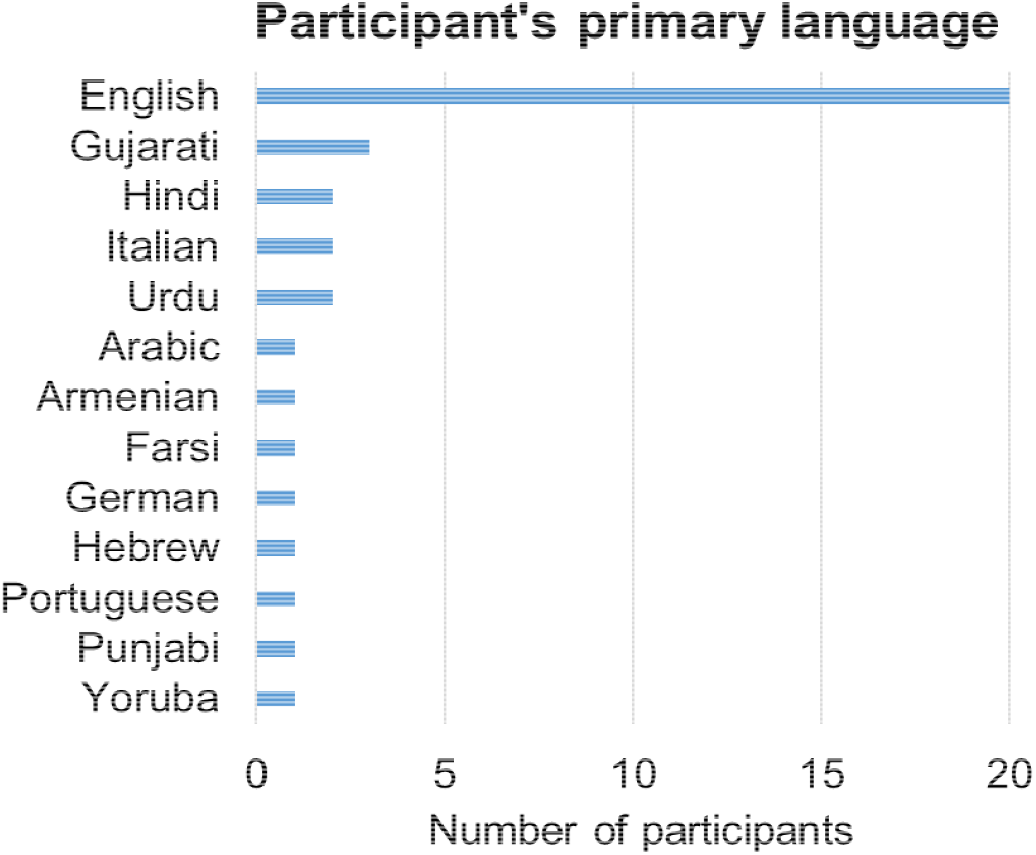
Primary language(s) spoken, as self-reported by participants (n=30)

Better eye median (IQR) logMAR visual acuity (VA) was 0.30 (0.17, 0.58). Nineteen (63%) of the 30 participants had prior experience of intravitreal injections, while 11 (37%) were injection-naïve. Eleven (37%) of participants had centre-involving GA.

When asked to self-report their GA severity (Appendix 1, Q16), 13 participants self-rated their GA as mild, 13 as moderate, and 4 as severe. A more severe self-report was associated with worse VA in the better eye (*r*_*s*_ (28) *=* 0.40, *p =* 0.029). This is consistent with previous reports demonstrating that vision-related quality of life is primarily dependent on the better eye.^41^ However, there was no statistically significant correlation between self-reported GA severity and: worse eye VA; VA in the GA eye; VA in the fellow eye; GA laterality; or centre-involvement.

Median (IQR) time to travel to the eye clinic was 30 (15, 45) minutes. As shown in Figure 3, 10 (33%) participants lived alone while the other 20 (67%) lived with spouses or partners, children or carers.

**Figure 3.**
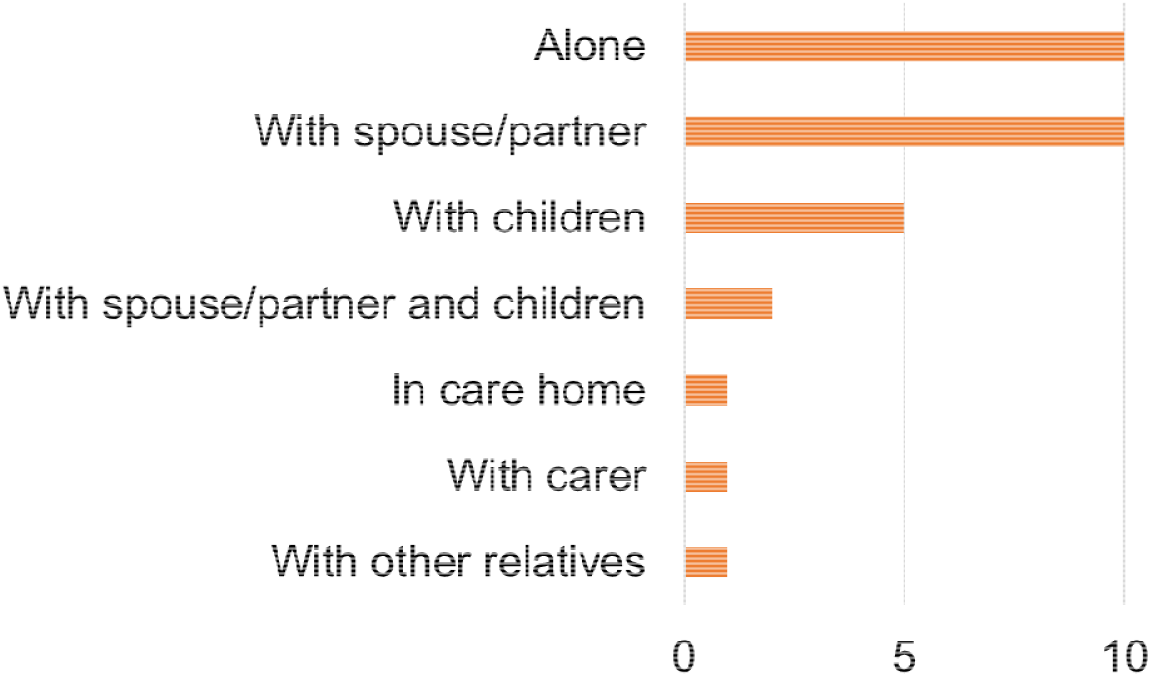
Living circumstances of participants (n=30)

Fourteen (47%) participants reported attending eye clinic appointments alone, while the other 16 (53%) were accompanied by relatives, friends or caregivers. Twenty-three (77%) of participants reported living with other chronic health conditions apart from AMD/GA, with 8 (27%) living with diabetes. In the EQ-5D, the domains in which participants reported most problems were mobility (mean score = 2.3) and usual activities (mean score = 2.1).

Interview times with participants ranged from 27 minutes to 120 minutes. Twenty-four of the interviews (80%) were conducted in person, and six (20%) by telephone.

### Quantitative findings on acceptability of intravitreal injections for GA

Findings from the Likert-type scale questions about acceptability of GA treatment are shown below in Table 3, while Figure 4 displays responses to questions about participants’ willingness to undergo intravitreal injections at different intervals. Figure 4 demonstrates the increase in acceptability when injections were proposed every other month rather than monthly, with 15 of 30 (50%) participants extremely likely to accept GA injections every other month, compared with 9 of 30 (30%) extremely likely to accept monthly GA injections.

**Table 2.** Responses to Likert-type scale questions on acceptability of GA treatments

| Likert-type scale question and responses | *N* | % |
| --- | --- | --- |
| **In your view, are the risks of the injection procedure, as explained, worth the potential benefit of slowing down the progression of geographic atrophy?** |  |  |
| Yes | 17 | 57 |
| Not sure | 11 | 37 |
| No | 2 | 7 |
| **Are you afraid of having an injection in your eye?** |  |  |
| Yes | 10 | 33 |
| Not sure | 4 | 13 |
| No | 16 | 53 |
| **Are you concerned about the side effects of injections into your eye?** |  |  |
| Yes | 10 | 33 |
| Not sure | 3 | 10 |
| No | 17 | 57 |

**Table 3.**
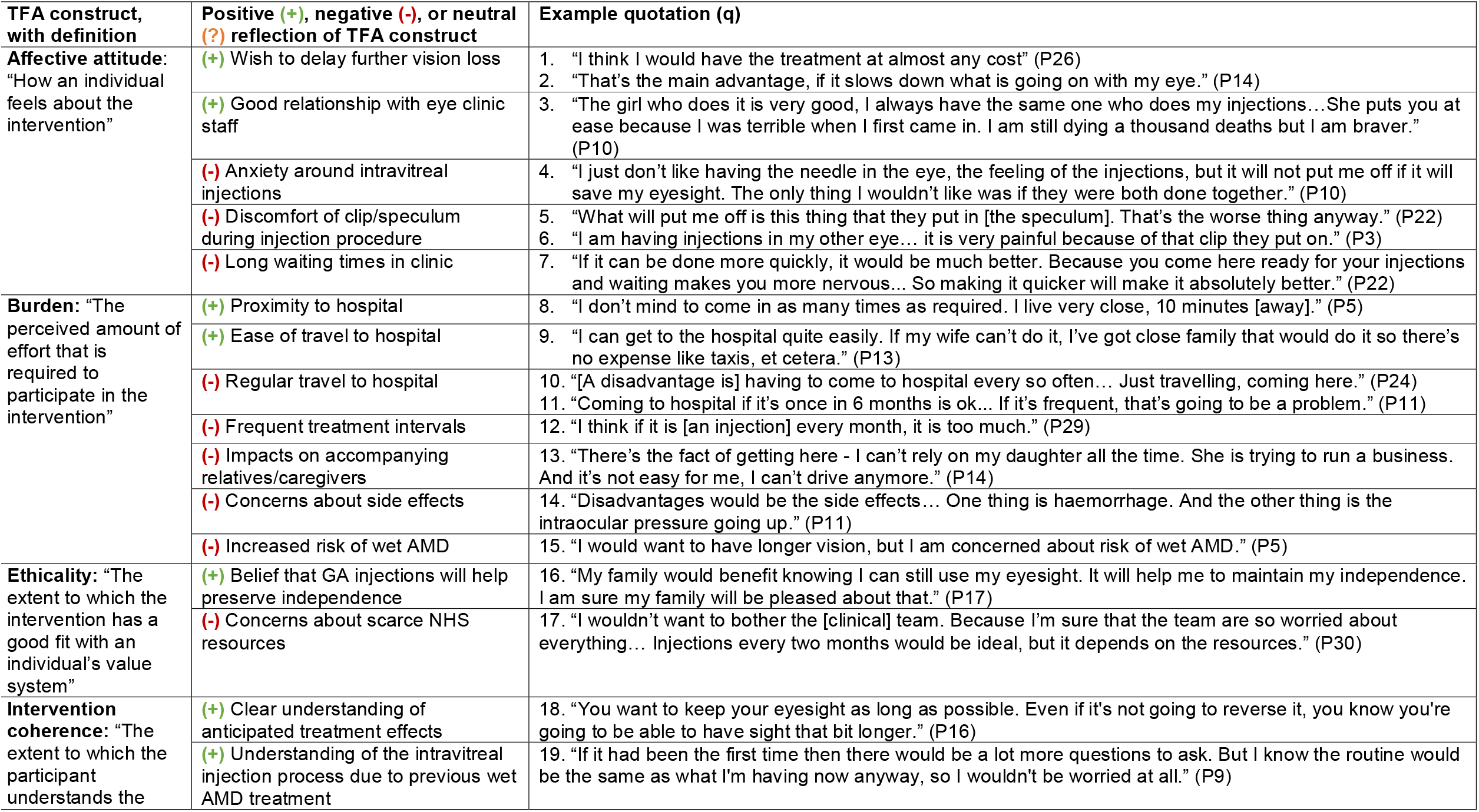
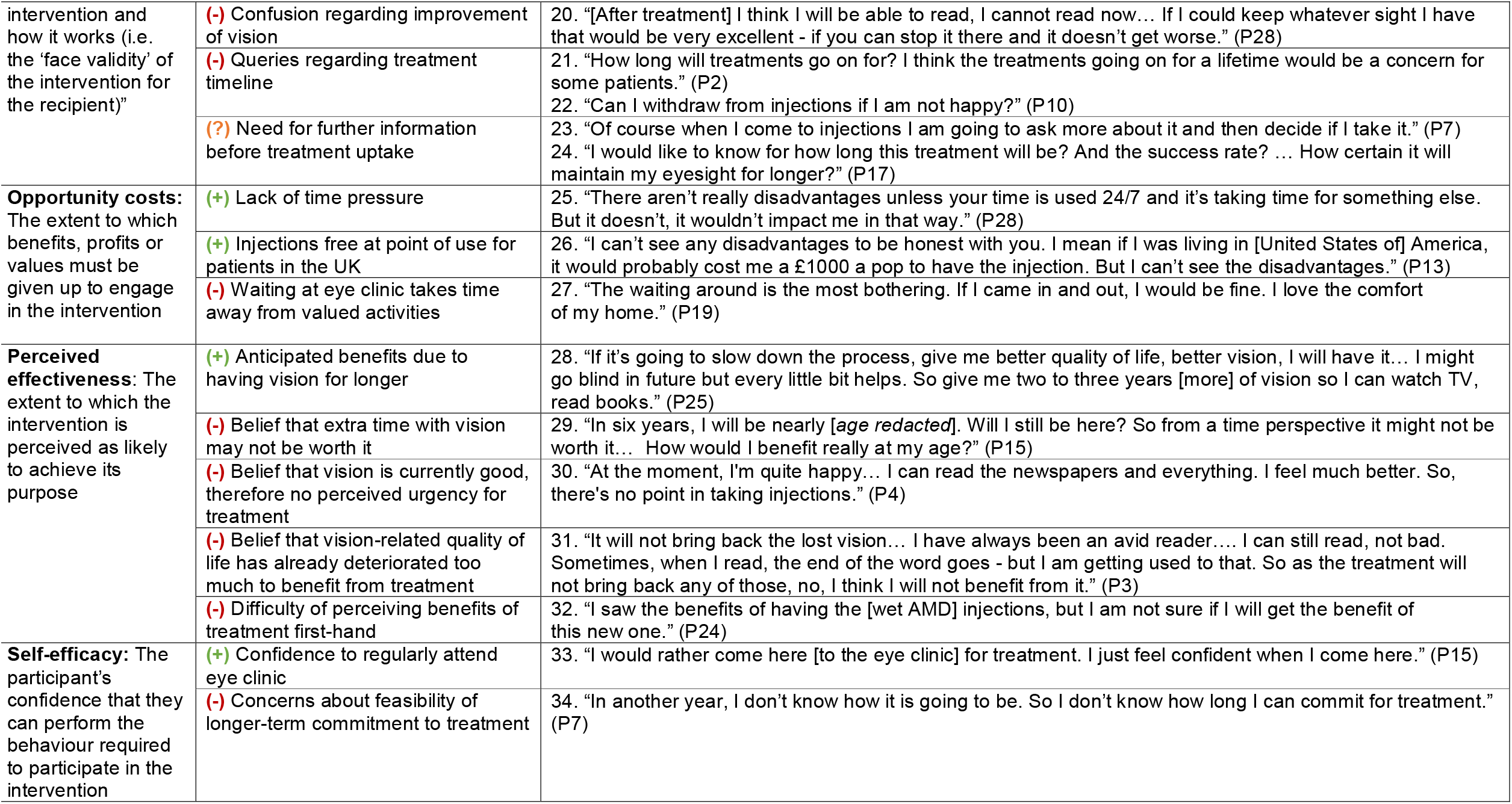
Participant reflections on prospective acceptability of GA treatment, categorised within the seven component constructs of the TFA

**Figure 4.**
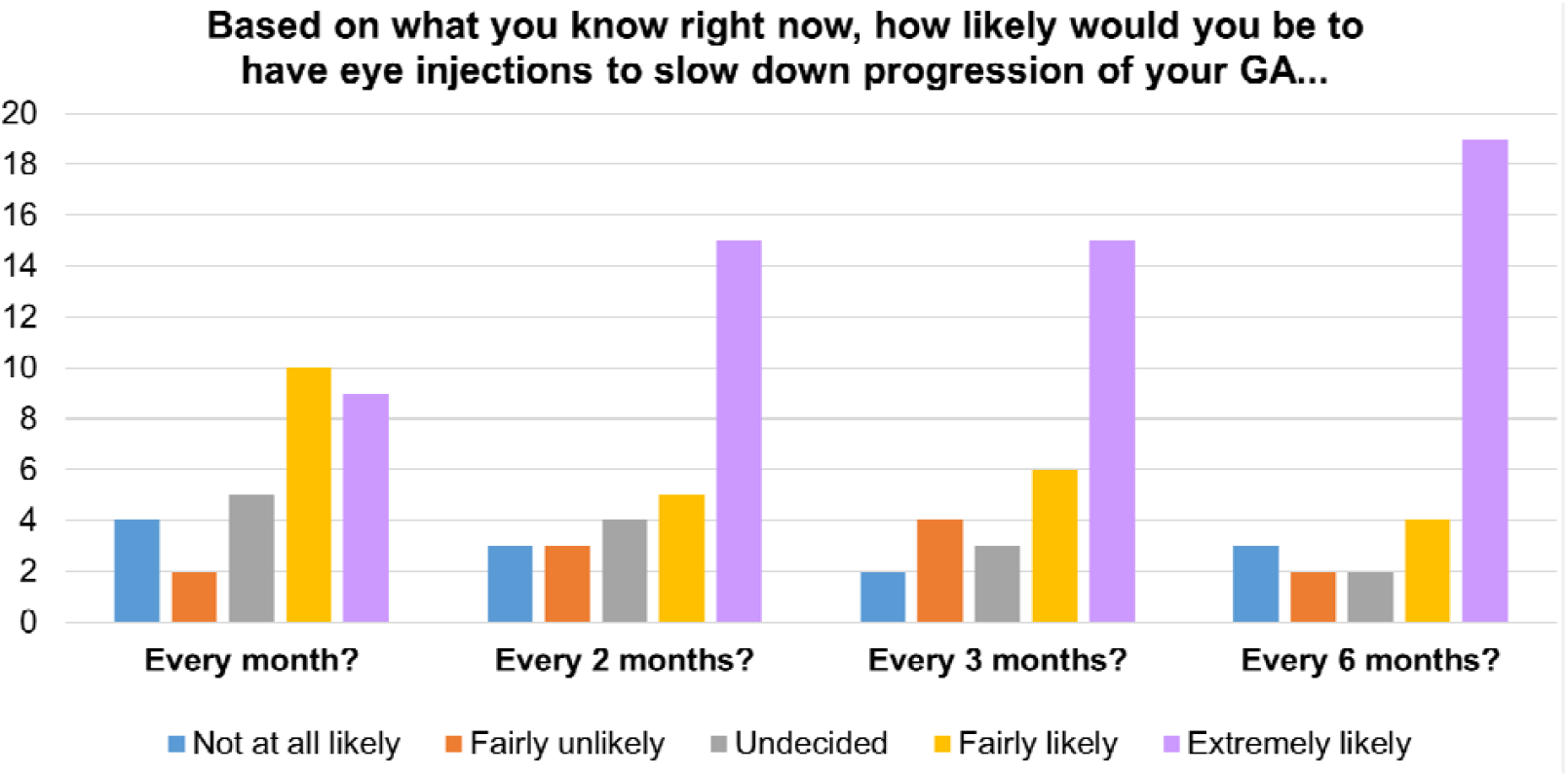
Responses to questions on acceptability of GA treatment at different intervals

Qualitative responses analysed within the TFA (see below) were additionally categorised into three groups, following analysis of the qualitative framework and reaching consensus among all authors. Eighteen (60% (95% CI: 41-79%)) participants were deemed to be positively accepting of the treatment despite their awareness of the burdens and drawbacks, and this group was termed “Treatment at any cost”. Eight (27% (95% CI: 10-43%)) participants were deemed to be “Ambivalent”, hesitant about treatment and unsure about the balance of benefits versus risks and drawbacks. Four (13% (95% CI: 0-26%)) participants were deemed “Unlikely to proceed” with treatment. These figures correlate strongly with participants’ responses on the Likert-type scale question asking whether the risks of treatment are worth the benefits (Table 2), *r*_*s*_ (28) = 0.69, *p* < 0.001.

Inferential analysis demonstrated a statistically significant, moderate correlation between overall acceptability level (i.e. membership in the three groups discussed in the paragraph above) and EQ-5D score, *r*_*s*_ (28) = 0.42, *p* = 0.021. Participants with worse self-reported health (higher EQ-5D score) were more likely to be in the “Treatment at any cost” group.

When considering correlations between other Likert-type scale question responses and demographic/clinical factors, statistically significant moderate correlations were only found for the question around concern about side effects of injections (Table 2). Concern about side effects correlated positively with: increased age, *r*_*s*_ (28) = 0.44, *p* = 0.014; presence of other chronic health conditions, *r*_*s*_ (28) = 0.47, *p* = 0.009; and naivety to intravitreal injections, *r*_*s*_ (28) = 0.43, *p* = 0.018.

### Qualitative findings on acceptability of intravitreal injections for GA, based around the Theoretical Framework of Acceptability (TFA)

Participants’ responses to the semi-structured, open-ended interview questions were coded into the seven constructs of the TFA.^28^ Table 3 displays the seven constructs as defined in the TFA, and different reflections of the construct as generated from participants’ responses, illustrated with example verbatim quotations. Quotations are subsequently referenced in the Results by number (e.g. q1 to denote quote 1 in Table 3).

#### Affective attitude

As shown in Table 3 (q1-2), positive affective attitudes towards GA treatment related to participants’ profound wish to delay further vision loss in the absence of other therapeutic interventions:

> “*If I take the injections, it will slow down the progression. I don’t mind about [it] being life long*.” (P12)
>
> “*I would do everything to have my eyesight for longer… The fact that there is any treatment, it’s a hope for people*.” (P17)

For some, a positive attitude to treatments related to their ability to slow deterioration while waiting for a more efficacious therapy (“*something could come along that would really reverse it*” (P9)), and also as a means of remaining within a secondary eye care pathway and thus potentially facilitating early detection of other eye diseases:

> “*We know [the treatment] doesn’t cure it but if it slows it down, and then obviously while they’re looking in the back of your eye they can find other things*.” (P13)

Several participants referenced their good relationship with clinic staff as a factor influencing their positive attitude to treatment (q3), that could reduce their anxiety around injections:

> “*To have a confident, experienced person doing it, is incredibly important. For the peace of mind of the person receiving it [the injection]*.” (P26)

More negative affective attitudes to the treatment often related to anxiety and concerns relating to the intravitreal injection procedure (q4), including among participants with experience of such injections for wet AMD:

> “*It is the pain definitely that scares me. I am very anxious even before the injection to a point that I don’t sleep the night before my appointment… If it could be eye drops, [that] would be much easier*.” (P17)

There were particular concerns around the discomfort of the speculum (“*I don’t mind the injections. It’s the clamp I don’t like*.*”* (P1) – and q5-6), and concerns around concurrent injections for GA and wet AMD (q4):

> “*I wouldn’t want the two done in one day at the same time. I think the pain would be too much with this clip*.” (P3)

While participants commonly reported anxiety about intravitreal injections, a significant minority – mostly, but not exclusively, with previous experience of intravitreal injections - did not see the procedure as a barrier:

> “*I don’t mind having injections, it is not painful. If we have to have injections, we are happy and we will come whenever you ask us to come*.” (P8)

Another key issue negatively influencing attitudes to the treatment related to the anticipated waiting times at the eye clinic, not only the time commitment in itself but also because waiting often intensified patients’ anxiety about the injection (q7). Participants accepted that the procedure would inevitably take time and acknowledged demand on ophthalmology services, but still hoped for an accelerated process:

> “*I know there is a lot to do and a lot of people having it done. You can’t just get it at the click of a finger. But quicker would be better*.” (P19)

At the same time, for some participants waiting time did not significantly influence their attitude to treatment:

> “*I don’t mind waiting for my turn to be seen. Also they haven’t made me wait longer than necessary*.” (P24).

#### Burden

Participants’ perspectives within the TFA Burden construct largely clustered around two main issues: firstly, the logistics of regular travel to the eye clinic, potentially alongside relatives, friends or caregivers; and secondly, side effects and the risk of developing wet AMD in the GA eye.

For some participants, their proximity to (q8) and/or ease of travel (q9) to the eye clinic meant that regularly attending injection appointments was perceived to require little effort. However, for others the regular, frequent travel to hospital for potentially lifelong injections was a burdensome prospect (q10-12), Some participants reflected that relatives or caregivers accompanying them to the eye clinic might also feel burdened (q13), although for other participants this was not always considered an imposition (“*My daughter is really good. She brings me to hospital for my appointments, she wouldn’t mind waiting. I suppose it gets on her nerves sometimes but she does it*” (P15)). Indeed, many participants reflected on how the effort required to attend injections might be differentially influenced by living situation, mobility, and/or transport options:

> *“Clearly, for some people it will be the cost. If you don’t have a lot of money then even the bus fare can be an issue. Mobility is a major issue*.*”* (P29)

One suggestion to mitigate the logistical burden was hospital-arranged transport: “*I think if you could organise transport to bring me to hospital it would be easier*.” (P29).

Participants were asked whether hypothetically they would prefer injections to be delivered at their local optometrist rather than coming to hospital; for some participants, that would reduce burden (“*Would my opticians do the injections? Oh that’s a good idea! It is a very good idea to have it in local close proximity*.” (P14)). However, other participants expressed a strong preference for injections at hospital, citing trust in their doctor and feeling safer in case anything went wrong: *“I would want to come to the hospital. Because it’s much safer, I feel safe*.” (P17)

Within the construct of Burden, participants also discussed side effects (q14) and the risk of developing wet AMD in the GA eye (q15). For some participants, concern about side effects was a major barrier to treatment acceptability (e.g. “*Side effects can hold me back from having injections*. (P21)), with several referring to their fear of “*something going wrong… The eye is a very delicate place”* (P29). However, other participants had few concerns about side effects of treatment, having become familiar with the intravitreal injection process through wet AMD treatment:

> “*All the stuff they put in there is the safeguard of all that infection-wise. So I don’t get worried about that now. The first couple of times I was always worried that it could get a little bit of a bleed, but that’s not a problem apparently*.” (P9)

Wet AMD was considered by some participants as “*the more dangerous one*” (P13), and this increased risk due to GA injections was described as “*off-putting*” (P25) and leaving participants “*concerned*” (P5, P10, P25). While there were widespread concerns about wet AMD, there were a small number of participants who did not see this as a concern, for example because they were already living with wet AMD and this was being successfully treated:

> “*I have the wet form in my other eye and I am having injections anyway, so having wet in this one [the GA eye] as well doesn’t make difference*.” (P6)

As discussed under Affective attitude, some participants were worried about the idea of two injections (for wet AMD and for GA) on the same visit. However, one participant expressed a preference for simultaneous injections which would allay the potential burden of developing wet AMD:

> “*The additional risk of getting wet AMD wouldn’t be an offput - because it wouldn’t be an extra session every month*” (P26).

#### Ethicality

The TFA construct of Ethicality, concerning the fit of the intervention with the participant’s value system, was somewhat less salient in participants’ accounts than other constructs. The only recurring theme within the Ethicality construct centred around norms of independence and self-reliance, valorisation of which would encourage many participants to take up GA treatment (q16). One participant also discussed concerns about overburdening healthcare staff and straining scarce resources were she to undergo frequent injections (q17).

#### Intervention coherence

There was a high degree of variation among our sample in terms of understanding the aims of treatment and its anticipated effects. Some participants clearly evidenced their understanding of the mechanism of GA intravitreal treatments being to slow down, rather than stop or reverse, deterioration thus enabling some retention of good vision for longer (q18). However, even participants who at first seemed to understand the treatment effects were essentially anticipating a relative “steadying” of vision loss (rather than continued, but slower, deterioration):

> *“I know it [the treatment] will not improve my vision, but as long as it will make it steady that’s better*.*”* (P2).
>
> *“If I didn’t have the injections, it may go a lot quicker. If [treatment] stabilises it – well, I know it will not stabilise it - but if it will hold it back, that would be perfect*.*” (P14)*
>
> “*If I know it will slow the progression and it stays as it is now, I don’t mind… I wouldn’t see any difference, would I?*” (P10)

These quotations highlight the complexity for participants in understanding the treatment effects, and the importance of accurately setting patient expectations before initiating treatment. As the third quotation (from P10) shows, some participants were aware they would not be able to perceive a noticeable difference in their GA progression first-hand, but still assumed their vision would remain at the same level. There was even lower intervention coherence among participants who believed the treatment could reverse their vision loss (q20), for example restoring their ability to engage in functional activities such as reading (“*I will be able to see longer and read. I can’t read well now*” (P6)).

Common misunderstandings or queries also centred around the potentially lifelong timeline of treatment (q21-22), with several participants lacking understanding that the treatment would require long-term adherence and persistence to remain effective:

> “*I try the injections*… *If it’s working, I continue it. If it’s worse for the eye, I will stop*… *So if it is the same after one, I continue, same after two, I continue. But if it is not better after three [injections], I will stop*.” (P7)

In terms of the actual procedure, some participants with experience of intravitreal injections for wet AMD were clear that they knew what to expect and would therefore have few queries (q19). Several participants believed that they would need further information before initiating treatment (q23-24), particularly regarding guarantees of treatment efficacy.

#### Opportunity costs

Opportunity costs refer to the extent to which benefits, profits and values would be given up to undergo GA treatment. While many participants discussed a negative affective attitude towards long waiting times in the eye clinic (see ‘Affective Attitude’ above), only one participant (q27) articulated what she would be giving up in order to commit time to injections. Conversely, one participant was clear that time in clinic would not be taking away from her time to do other things (q25), and therefore there was no opportunity cost.

While financial implications were discussed in the context of transport (see ‘Burden’ above), cost was not a factor discussed by participants because NHS treatment in the UK is free at the point of delivery to patients. One participant explicitly stated that in other health systems, treatment costs could be a major disadvantage for patients (q26).

#### Perceived effectiveness

In this study, we considered perceived effectiveness as the extent to which participants perceived GA treatment as likely to achieve its purpose and result in tangible benefits. Often the benefits referred to by participants related to prolonging their participation in functional activities such as driving and reading, leisure and social and/or family activities (e.g. “*I’m hoping that I can continue playing golf for longer… As long as it keeps my vision as it is I’m well capable of driving, it’s not a problem at all. I drive and see my grandchildren*.” (P13)). However, some participants – even if overall accepting of the treatment – voiced disappointment that the treatments can only slow down, but not stop or reverse, GA progression: *“Well, er… that’s the best they can do? Slow down the process?”* (P30).

For many participants, the benefits of slowing down vision loss were presented in the context of their advanced age, meaning that even a modest amount of time with preserved vision would be valuable (q28). As one participant stated:

> *“[Treatment will allow me] to keep what I have got, to carry on doing things longer. As I said I am [86-90 years old], I don’t know how many years I’ve got, I will not live up to 200*.” (P10)

However, conversely, some participants referred to their age as a factor which would counteract the perceived effectiveness of treatment, unsure if the benefits outweigh the efforts and drawbacks of regular injections as they looked to the future (q29). For example, one participant with good visual acuity (0.16 logMAR) in her GA eye stated:

> *“I am [81-85] years old, if I have 2 more years I will be satisfied… I think I’ll go on like this for another couple of years*.*”* (P11).

Indeed, one participant (0.4 logMAR in GA eye, 0.2 logMAR in fellow eye) discussed his concerns around perceived effectiveness on the basis that he saw no rationale for injections (q30), because he perceives his current visual function as good and he does not want to risk “*my eyes being affected in any way*” (P4 – see OCT images in Appendix 4). This participant’s attitude is consistent with the notion that visual function is driven primarily by the better-seeing eye. However, another participant (0.04 logMAR in GA eye, 0.86 in fellow eye) voiced a contrasting perspective, namely that his good level of vision would enable him to regularly attend the eye clinic for GA treatment:

> *“I have good vision now, in my right eye, the left eye is wet. It does not affect me, I don’t mind coming for treatment as many times as needed*.*”* (P5)

At the other end of the spectrum were participants with advanced vision loss who saw the perceived effectiveness of treatment as low, since it could not restore their lost vision and thus improve their vision-related quality of life (q31). For example, one participant (CF in GA eye, 0.92 logMAR in fellow eye) stated:

> *“If there is no improvement in my life - for example I want to read, I can’t read my bible, that pains me - then what’s the point? It will put me off [treatment]*.” (P23)

Participants also discussed a more conceptual point, namely that patients being treated would not be able to see benefits first-hand in terms of their vision improving, in contrast to wet AMD treatment (q32). As another participant stated, *“If they keep on having the injection and they can’t see a benefit, they won’t be happy*.*”* (P23). One participant alluded to the fact that benefits of treatment would only be perceptible in the long-term, in the absence of any immediate effect after each injection:

> *“Well I wouldn’t like to do it for nothing, to get no results at all. But I wouldn’t know until it’s done*.” (P30).

#### Self-efficacy

Participants only sparingly discussed issues relating to Self-efficacy, their confidence in their ability to undergo the treatment. Comments within this construct related to confidence in the injection procedure being carried out in a hospital rather than optometrist setting (q33), and to uncertainty regarding their confidence in committing to lifelong injections (q34).

### Qualitative findings beyond the TFA

Themes were also generated inductively from aspects of participants’ accounts which fell outside the constructs of the TFA, but were still relevant to GA treatment acceptability. These themes and associated quotations are presented in Table 4.

**Table 4.** Themes relating to GA treatment acceptability outside the TFA constructs

| Theme derived from framework analysis | Description | Illustrative quotation(s) |
| --- | --- | --- |
| Inter-individual differences in GA treatment acceptability | Participants discussed their perceptions of how treatment acceptability might differ across patients because of various clinical or demographic factors or depending on previous intravitreal injection experiences. | 35. "A lot of it will come down to people's ages. You know, if you get somebody who's in their 80's, they might think, 'Well I can't get to the hospital once a month, I'd rather it be longer.'" (P9)<br>36. "I cannot speak for others. But it probably depends where they live, how far they come from, how many members of the family they need to get help. It depends on these things." (P2)<br>37. "It really depends on the age of the patient and also depends on condition of their fellow eye... It's like [the] Covid vaccine, it's an individual decision." (P11)<br>38. "I suppose if you've got the dry one, you would probably want it. But if you have already wet in one eye, it's hard to say whether you want [injections] in that eye as well." (P15)<br>39. "It's better to get something done. You see, because I had all those injections, so many in this eye, I know how it feels and how it happens. That's why I don't mind having injections." (P2) |
| Trust in clinicians' professional judgement about the value of treatment | Several participants suggested that their decision to undergo treatment would ultimately be based on their doctor's professional | 40. "There is nothing that would put me off. The doctors know better. If they think I need it, I am happy to come for injections." (P5)<br>41. "If doctors say it will help me, I don't mind having injections." (P8)<br>42. "I am not so sure about the benefits. But I trust my doctor, trust my hospital." (P22)<br>43. "I'll agree with whatever they think is the best for me. Whatever the experts think is the best for me, I'll go with." (P13) |
|  | judgement of whether they would benefit. |  |
| Longer term hopes and wishes for treatment | Participants discussed their hopes in the longer term that a more efficacious treatment could be found to improve their vision. | 44. ““If it [intravitreal treatment] stabilises it [GA] until something else can be found that will retract it back a bit, that would be the thing to look forward to...” (P9)<br>45. “I really hope they will find treatment in future for me, for people. I hope it will improve the vision.” (P5)<br>46. “At my age I don’t think I’ll get any benefit. But for future generations I am glad that there is a hope.” (P24) |
| Impact of GA on quality of life | Participants discussed the wide variety of the functional and emotional impacts of GA, which were often closely intertwined. Participants also discussed how the impacts of vision loss also affected family members. | 47. “I’ve been acutely aware over the last year and a half how my eyesight for reading has deteriorated... I found that incredibly frustrating and incredibly upsetting.” (P26)<br>48. “I can’t rely on my eyesight anymore. It’s restricted, I suppose, certain things for me. There are lots of different things that I can’t do now that I used to happily do. I used to do a lot of cooking and I am not doing it now.” (P14)<br>49. “Of course if I can get treatment my family will be happy... I could do things in the house but now I am not able to. So if I have treatment everybody will be happy. It affects my wife because she sees me down and unhappy sometimes.” (P2)<br>50. “I don’t think it affects me emotionally because I can’t get even more anxious than I am. So to me, it is one of those things you just accept.” (P3) |

## Discussion

Our study findings suggest that a majority of GA patients would be accepting of intravitreal treatment for GA, whilst recognising potential burdens and inconveniences. The key concern for people with GA, which emerged in our study as the central underpinning motivation for treatment, is the high priority of continuation with vision-specific activities, particularly for those in worse self-reported health. For 60% of the study participants, despite acknowledging potential drawbacks, the possibility of extending the time they have to engage in vision-specific activities and remain independent was deemed a worthy trade-off, and they would therefore opt for ‘treatment at any cost’.

The factors limiting acceptability were largely clustered around concerns about magnitude of treatment efficacy, fear of wet AMD and side effects (and to a lesser extent, the injection procedure itself), and logistics of regular eye clinic visits for treatment. Specifically, reducing the frequency of injections from monthly to every other month increased the proportion of participants that were extremely likely to accept these treatments if offered now.

Factors maximising acceptability were the wish to continue with vision-related activities for longer, and the hope that these treatments might slow GA progression until more efficacious therapies emerge. Participants who self-reported a worse state of health at the time of the study (as measured by the EQ-5D) evaluated the treatments as significantly more acceptable.

Crucially, as discussed within the TFA’s Perceived Effectiveness construct, there were a number of participants with better visual acuity than the sample average who saw no value in treatment, because they perceived their vision as currently good and thus saw no rationale for treatment. However, natural history studies demonstrate a progressive decline in vision over time, with almost two-thirds of eyes observed to have foveal involvement associated with moderate or severe sight loss within 4-5 years.^27,42^ Additionally, the current treatments have been demonstrated to have higher efficacy the further the lesion is from the fovea,^43,44^ thus extending time of foveal preservation. Therefore, there is a challenge here to accurately identify and robustly support patients at risk of foveal involvement in future whilst their visual acuity remains good, in order to maximise potential to preserve vision with these treatments if approved.

Given the heterogeneity of GA in terms of progression, observation of recent progression over time with multi-modal retinal imaging could be a useful way to demonstrate the potential likelihood for the individual patient to benefit from these treatments. Further work is required to develop precise and robust risk stratification tools and to determine the time-difference in progression that patients will perceive as meaningful. Data from Colijn and colleagues’ analysis of four population-based cohort studies^27^ suggests that delaying progression to foveal involvement by at least 0.8 years could allow the average individual with non-foveal GA to retain central vision and avoid severe vision loss for the rest of their life.^45^ As such, even a modest reduction in rate of progression would deliver clinically meaningful benefits to a large number of patients.

Listening to our study’s participants, it is vital that patients are effectively counselled, and educated on the natural history of GA and accurate expectations of treatment effects; including the fact that they are unlikely to perceive treatment benefits directly, and can expect their GA to continue to progress, albeit at a slower pace. Treatment initiation should follow a shared decision-making process involving the patient and their eye care team.^46,47^ Since participants also noted that their stance on treatment may change over time, counselling on treatment expectations should take place regularly to support adherence,^21^ rather than simply being a one-off event.

Within the Burden construct, the increased acceptance of every other month injections is worth highlighting, particularly given recent 24-month outcome data from the DERBY and OAKS phase 3 registration trials. These trials demonstrate a marginal difference in GA growth reduction between the monthly and every other month treatment regimen (19% reduction for eyes treated monthly vs 16% reduction for eyes treated every other month in DERBY; 22% reduction for eyes treated monthly vs 18% reduction for eyes treated every other month in OAKS).^48^ On the other hand, monthly injections in these trials were associated with a near doubling of the rate of exudative choroidal neovascularisation (11.9% in monthly versus 6.7% when treated every other month). Similar rates of choroidal neovascularisation have been reported in the avacincaptad pegol trials.^49^ An every-other-month regime could thus deliver increased adherence and persistence, a better safety profile (almost 50% reduction in neovascularisation risk) and greater cost-effectiveness for healthcare funders, with only a minimal reduction in efficacy.

Furthermore, participants’ fear of wet AMD risk commonly emerged as an off-putting aspect of treatment, although for some participants this was less of a concern because of the availability of a more efficacious treatment for wet AMD, or if they were already being treated for wet AMD. Even for study participants generally accepting of the GA treatment, the prospect of injections on the same day for wet AMD and for GA was burdensome (although there was one participant – P26 – who welcomed the convenience of consecutive same-day injections). Given that a 2-3 fold increased risk of wet AMD as demonstrated in the phase 3 trials^44,49^ may necessitate regular monitoring and retinal imaging for these patients and increased costs to payers, there may be an urgent need for innovative service delivery to support the rollout of these treatments if approved. Shared-care models involving monitoring by community optometrists may help expand capacity and reduce time spent in hospital clinics. Our results confirm that longer-acting therapies which slow progression to a higher degree or even halt atrophy remain an unmet need, and must be the focus for future drug development. In the meantime, more frequent review of their ocular health may well be welcomed by many GA patients, who are typically discharged from eye clinics in the UK at present, with no targeted psychosocial support for what is a progressive and debilitating disease.^12,14^

### Strengths and Limitations

Initially conceived as an exploratory pilot study, our study has a number of limitations. Firstly, as a relatively small-scale study involving patients from two London-based sites, there is limited generalisability to other contexts, for example other geographies in the UK (e.g. rural populations) or other countries with different eye care systems. Secondly, our system of categorisation of participants into three acceptability groups was undertaken in response to emergent patterns in our framework matrix, but did not follow a standardised method that had been predetermined in our protocol. This categorisation could variously be considered too subjective or reductive, and our forthcoming larger, multi-site quantitative study will provide a more robust, generalisable quantification of GA treatment acceptability. Thirdly, while the TFA was used to analyse the data, our interview topic guide was not systematically developed from the TFA; instead, more open-ended questions were used to explore participants’ hopes, beliefs and concerns around treatment, based on our literature review and the insights of our study’s patient advisory group. This meant that for certain TFA constructs (e.g. Ethicality and Self-efficacy), there was less rich discussion than there may have been, had the TFA been used expressly to shape the topic guide.

Nonetheless, in terms of strengths, this is the first study systematically exploring prospective acceptability of GA intravitreal therapy among a diverse sample of patients, recruited using maximum variation sampling to try to ensure participants were representative of the broader GA population. While conceived as a qualitatively-driven mixed methods study,^50,51^ the quantitative element helps to corroborate and (tentatively) quantify interpretations made on the basis of the qualitative data; indeed, there was close alignment between responses to the Likert-type scale questions and patterns in the qualitative data. Analysis of the qualitative data using the robust Theoretical Framework of Acceptability allowed us to make sense of a rich and complex dataset, and to identify the key motivating factors driving acceptability and what most concerns GA patients and could be modified in future.

## Conclusion

In summary, a majority of participants (∼60%) were positive about GA treatment, despite the potential inconvenience and burdens, largely because slowed progression could allow for continuation with vision-specific activities and because current treatments may slow GA until more efficacious therapies become available. Indeed, our study findings suggest that the extent of GA’s impact on patients’ vision-related quality-of-life would strongly influence their choice to take up (or forgo) a potentially burdensome treatment. Participants’ key concerns related to the modest efficacy of treatment, the risk of wet AMD and side effects, and logistical issues associated with frequent, potentially lifelong treatment.

There is also a sharp increase in patient acceptability when considering an every-other-month treatment regimen in comparison to monthly treatment. Given encouraging efficacy and safety outcomes for the every-other-month regimen, this may be an optimal dosing label for patients, payers and health services.

The findings also highlighted issues in the Intervention Coherence domain, with frequent misunderstandings regarding the workings and likely effects of the intravitreal treatments, which demonstrates a need for clear, accessible patient education tools should emerging intravitreal therapies be approved.

Further research to understand the patient perspective in a larger population of patients with GA is required to confirm our findings, and identify any correlations between patient acceptability and structural and functional biomarkers of GA severity. We expect such research to aid patient education, selection and individualisation of treatment regimes.

## Supporting information

Appendix 1 - Study questionnaire

Appendix 2 - Codebook for framework analysis

Appendix 4 - OCT images for Participant 4

## Data Availability

The most pertinent data produced in the present work are contained in the manuscript. The full interview transcripts generated during and analyzed during the current study are not publicly available, because the in-depth and specific information they contain could compromise the privacy of participants.

## Abbreviations and Acronyms

AMD: Age-related Macular Degeneration
GA: Geographic Atrophy
NHS: National Health Service
TFA: Theoretical Framework of Acceptability
VEGF: Vascular Endothelial Growth Factor

**Appendix 1: Study questionnaire**

**Appendix 2: Codebook for coding qualitative data**

**Appendix 3: Demographic and clinical characteristics of study participants**

**Appendix 4: OCT images**

