## Appendix 1 - Study questionnaire for "Exploring patient acceptability of emerging intravitreal therapies for Geographic Atrophy: a mixed-methods study"

**Appendix 1: Telephone questionnaire**

### A. Socio-demographic questions

1. How old are you?
2. What is your ethnicity?

- **Options:** Black; East Asian; South Asian; White; Other (please specify)?

3. What is the primary language you speak?
4. What is the highest level of education you have achieved?

- **Options:** Primary; Secondary; University; Postgraduate; Other (please specify)?

5. What is your work status?

- **Options:** Retired; Unemployed; Employed; Self-employed; Other (please specify)?

6. Do you usually have to take time off work to attend hospital appointments?
7. What is your current living situation?

- **Options:** Living with spouse/partner; living with spouse/partner and child(ren); living alone; living with other relatives; Other (please specify)?

8. How long does it take you to get to your eye appointment?
9. How do you get to your hospital appointment?

- **Options:** Public transport; Taxi; Driven by family/friend or carer; Walk; Drive (self); Other (please specify)?

10. Do you attend your hospital appointments alone or does someone usually come with you? If someone comes with you, who are they?
11. Do you have any other long-term health conditions apart from AMD/GA? (If yes, ask participant to specify)
12. Approximately how often would you typically attend hospital appointments in an average year (for example, 2019), for reasons other than your AMD/GA?
    - **Options:** Once per year; twice per year; three times per year; more than three times per year?
13. Have you ever had injections in your eye? (**Options:** Yes/No)

13a. [*If Yes*] How many eye injections have you had?

- **Options:** Less than five; five to 10; 11-15; 16-20; More than 20?

13b. [*If No*] Has anyone close to you (e.g. a spouse, partner, other relative etc) ever had eye injections? (**Options:** Yes/No)

### B. Quality of Life questionnaire – EQ5D, 5 level version

[Redacted for copyright. EQ5D questionnaire is available through EuroQOL - <https://euroqol.org/> ]

### C. Pre-information structured questions (Likert-type scale)

| **14.**  How well do you feel you understand your geographic atrophy? | 1  Don’t understand at all | 2  Understand slightly | 3  Understand moderately well | 4  Understand very well | 5  Understand extremely well |
| --- | --- | --- | --- | --- | --- |
| **15.** How concerned are you about your geographic atrophy? | 1  Not at all concerned | 2  Slightly concerned | 3  Moderately concerned | 4  Very concerned | 5  Extremely concerned |
| **16.** How would you grade your geographic atrophy? | 1  Not bad (Mild) | 2  Bad (Moderate) | 3  Very bad (Severe) |  |  |
| **17.** How long do you think your GA will continue? | 1  A very short time | 2  A short amount of time | 3  A moderate amount of time | 4  A long time | 5  Forever |
| **18.** Based on what you know right now, would you consider eye injection treatment to be a good or bad development for geographic atrophy? | 1  Very bad | 2  Fairly bad | 3  Undecided – neither good nor bad | 4  Fairly good | 5  Very good |

### D. Re-iteration of information about treatments

*Thank you for answering all these questions. We are now going to provide more information about the treatments for Geographic Atrophy currently being tested out in clinical trials. These trials are happening completely independently, and have no relation to the research interview that we are conducting with you now.*

- *Currently, there are three promising injection treatments in the final stages of testing.*
- *These treatments have all been found to be safe. Clinical trials are now being carried out to determine how effective these treatments are in slowing down Geographic Atrophy.*
- *These treatments are all delivered by injection into the eye.*
- *The key point to note is that these treatments will not cure, stop or reverse Geographic Atrophy. Instead, they will slow down the vision loss caused by Geographic Atrophy.*
- *It is estimated that these treatments can slow down vision loss by up to 30%. As a concrete example: without treatment, a person could be five years away from having to stop watching TV because of Geographic Atrophy. However, if they were having the treatment, then they could continue to watch TV for up to eighteen months longer.*
- *For the treatment to keep working, it would involve likely lifelong, regular visits to the hospital for an assessment, followed by injection into the affected eye. These visits could be every month, every other month, or every three months depending on the exact treatment. However, it is likely that the less frequent treatments may have slightly less benefit.*
- *Injections into the eye may cause mild pain and discomfort in some cases, but severe side effects are rare. Early studies of some of these treatments have shown a small increase in the chance of developing wet AMD in the same eye as the dry AMD, in patients treated with the new treatments compared to patients with GA who did not have the new treatments.*
- *Eye injections are the standard of care for wet AMD, and used very frequently and safely to prevent vision loss from wet AMD. Over 400,000 eye injections are delivered every year in the UK with an excellent safety record.*
- *Injections in the eye can cause anxiety. However, people often feel much less anxious after having the first injection. Drops are used before these injections, in order to numb the eye and minimise any pain or discomfort.*
- *Do you have questions on any aspect of this information?*

### E. Post-information structured questions

| **19.**  Are you afraid of having an injection in your eye? | 1  Yes | 2  No | 3  Not sure |
| --- | --- | --- | --- |
| **20.** Are you concerned about the side effects of injections into your eye? | 1  Yes | 2  No | 3  Not sure |
| **21.** In your view, are the risks of the injection procedure, as explained, worth the potential benefit of slowing down the progression of geographic atrophy? | 1  Yes | 2  No | 3  Not sure |

| **22.** Based on what you know right now, how likely would you be to have eye injections ***monthly*** to slow down the progression of your geographic atrophy, if offered tomorrow? | 1  Not at all likely | 2  Fairly unlikely | 3  Undecided | 4  Fairly likely | 5  Extremely likely |
| --- | --- | --- | --- | --- | --- |
| **23.** Based on what you know right now, how likely would you be to have eye injections ***every other month***, to slow down the progression of your geographic atrophy, if offered tomorrow? | 1  Not at all likely | 2  Fairly unlikely | 3  Undecided | 4  Fairly likely | 5  Extremely likely |
| **24.** Based on what you know right now, how likely would you be to have eye injections ***every three months***, to slow down the progression of your geographic atrophy, if offered tomorrow? | 1  Not at all likely | 2  Fairly unlikely | 3  Undecided | 4  Fairly likely | 5  Extremely likely |
| **25.** Based on what you know right now, how likely would you be to have eye injections ***every six months***, to slow down the progression of your geographic atrophy, if offered tomorrow? | 1  Not at all likely | 2  Fairly unlikely | 3  Undecided | 4  Fairly likely | 5  Extremely likely |
| **26.** How do you feel about having a monitoring visit (i.e. an eye check-up) every time you have an eye injection? | 1  Very happy | 2  Quite happy | 3  Undecided – neither happy nor unhappy | 4  Quite unhappy | 5  Very unhappy |

### F. Post-information semi-structured questions

27. Specifically regarding the eye injection, is there anything that could put you off having the injection? (*Possible prompts: the process itself; the uncertainty of the benefit; the possibility of pain; the possibility of side-effects; the frequency of injections; the indefinite nature of the treatment…?*)

28. Does the fact that treatment will *slow down* vision loss*,* but not improve vision, put you off the idea of committing to long-term injections?

- *Prompt: Although the injections will not bring back vision that is already lost, are you willing to have injections to improve the possibility of doing your daily activities/hobbies for longer?*

29. What do you think would be the main benefit to you of the treatments we have described to you?

30. What do you think would be the main benefit for other people with geographic atrophy?

- *Prompt: Do you think other patients would think about the benefits in the same way as you?*

31. And what do you think would be the main advantages for people in your life (i.e. family/partner/friend who supports you at appointments)?

32. What do you think would be the main disadvantages to you of the treatments we have described to you?

33. What do you think would be the main disadvantages for other people with geographic atrophy?

- *Prompt: Do you think other patients would think about the disadvantages in the same way as you?*

34. And what do you think would be the main disadvantages for people in your life (i.e. family/partner/friend who supports you at appointments)?

35. How would you feel about these injections being delivered outside a hospital setting? For example, by a specially trained optometrist in a community setting? What might be the advantages and disadvantages?

36. What supports might help you to cope with the challenges of having these treatments? What could be done to make the injections as comfortable and convenient as possible?

37. What further information (if any) would be helpful to make your decision about whether to have these treatments?

### G. Discrete Choice Experiment task

[Focus of future paper]

### H. Final questions/wrap up

38. How does living with GA affect your everyday life? (*Prompts: How does GA affect your quality of life? Or does it?*)

39. How does living with GA affect your family members and/or friends? (*Or does it?*)

40. How does your geographic atrophy affect you emotionally *(e.g. does it make you angry, scared, upset or depressed*)?

41. What are the most important outcomes of treatment for you? What would you hope a treatment for GA could achieve?

42. How did you find answering these questions? Did they bring up any thoughts or concerns?
