## Appendix 2 - Codebook for framework analysis for "Exploring patient acceptability of emerging intravitreal therapies for Geographic Atrophy: a mixed-methods study"

**Appendix 2: Working codebook for AGAIN pilot, Version 4**

| **Category (with TFA definition where relevant)** | **Code** | **Example quote** |
| --- | --- | --- |
| ***Codes developed from Sekhon et al’s TFA*** | | |
| **Affective attitude**: “How an individual feels about the intervention” | Positive thoughts regarding treatment | “I think I would have the treatment at almost any cost”  “I don’t like to go to hospital all the time. I would like to do it locally.” (P1)  “If the vision I've got now is more or less going to stay the same, at that level, then yes, it would be nice.” (P9) |
|  | Ambivalence towards treatment | “Having injections sounds like a good idea. So whatever I do I should have something I could see out of. And if I have injections I wouldn’t want that to go. I really want to have some form of vision. If injections could destroy that, I wouldn’t want to go ahead. I am very concerned about problems with injections.” (P14)  “If I can see better it will be wonderful for everybody, for my family, for myself. But I hope to achieve something anyway, I hope. Because there is always risk. I’ll try, it can’t be worse anyway, can be? But I hope I will not have injections in the left one as well. Because [if] there are the risks from injections, I would rather avoid it. If I could avoid injections I would, believe me. I hope it will not get worse, I am not looking forward to injections.” (P22) |
|  | Attitudes towards injection procedure [including injection pain or discomfort] | “My mum says she still feels the pain. And speaking to a couple of ladies, they don’t feel it at all. That’s what bugs her sometimes. So if it’s not painful that would help. Feeling less of the injection would be relief.” (P25)  “The main anxiety is from injections. And being here as well. Hours of being here. She would prefer not being here. I have to encourage her.” (P25) |
|  | Attitudes towards time in clinic | “More comfortable if it can be done more quickly, it would be much better. Because you come here ready for your injections and wait makes you more nervous ’When is going to be my injection?’. You know, you get panic. I get really really anxious. So making it quicker will make it absolutely better.” (P22) |
|  | Familiarity/relationship with health professionals [as a factor affecting feelings towards treatment] | “The girl who does it is very good, I always have the same one who does my injections. I can’t think of anything that can be done differently, she is very good. She puts you at ease because I was terrible when I first came in. I am still dying thousand deaths but I am braver. The only thing I don’t like is the clip that they put on, it gives me more pain later than I have with injection. But when she knows it hurts she is very careful when she puts that in so that it’s comfortable.” (P10) |
| **Burden**: “The perceived amount of effort that is required to participate in the intervention” | Side effects and wet AMD risk | “I would kind of like to know about the side effects. Side effects can hold me back from having injections.” (P21) |
|  | Transport and logistics | “Well fortunately I can get to the hospital quite easily. If my wife can’t do it, I’ve got close family that would do it so there’s no expense like taxis, et cetera. I suppose getting to the hospital is the main thing and disadvantage to other people. Like I mean I’ve waited – I think the longest I’ve ever waited is two hours which is – so you expect that sometimes, little blips, but I can’t see any disadvantages to them other than getting to and from the hospital.” (P13)  “It’s not too long to come to clinic, I live couple of miles away, 15-20 minutes.” (P2)  “Regarding having injections in community setting: “That would be good of course. Because it’s a long way to come here [to the hospital]. Since 12 o’clock I am here today and wait, wait, wait. So if there is chance I would prefer [that], I don’t know if there is a chance.” (P22)  “So my opticians do the injections? Oh that’s a good idea! It is a very good idea to have it in local close proximity to me. So I will not need to come here [to the hospital], I can get my neighbour to drop me off instead of my daughter coming over.” (P14) |
|  | Impact on accompanying relative or caregiver | “Well it helps there will be no disadvantages. There will be none for me but probably for my son, because he always comes with me to hospital and wastes his time. I don’t mind waiting but he really doesn’t like it. It really bothers him.” (P6)  “Frequency of treatment is a real concern . They [my daughters] will have to take time to bring me to hospital. They don’t live with me and live far. Also they work, have families. I don’t want to bother them too much. If I could go to hospital myself, it would be good.” (P29) |
| Ethicality: “The extent to which the intervention has a good fit with an individual’s value system” | Independence, dependence, interdependence | “My family would benefit knowing I can still use my eyesight. It will help me to maintain my independence. I am sure my family will be pleased about that.” (P17) |
| Intervention coherence: “The extent to which the participant understands the intervention and how it works (the ‘face validity’ of the intervention for the recipient)”. | Patient queries and uncertainties [a rationale for more information and education] | “How long will treatments go on for? I think the treatments going on for a lifetime would be a concern for some patients.” (P2) |
|  | Patient understanding of the treatment | “I know it will not improve my vision, but as long as it will make it steady that’s better... I know vision doesn’t get better, but even if it will stay good for longer it is still good.” (P2)  “Realistically, my mum is [81-85 years old]. How many more years has she got? She takes 18 tablets a day. Realistically… what could she do at her age even if her vision was improved. It might not seem like a lot. But if she feels confident in a way that her vision is not going to deteriorate and she is not going to go blind that will mean a lot to her.” (P25) |
| Opportunity costs: “The extent to which benefits, profits or values must be given up to engage in the intervention” | Time | “The waiting around is the most bothering. If I came in and out I would be fine. I love the comfort of my home.” (P19) |
|  | Financial and employment impacts | “I can’t see any disadvantages to be honest with you. I mean if I was living in America, it would probably cost me a £1000 a pop to have the injection. But I can’t see the disadvantages.” (P13) |
| Perceived effectiveness: “The extent to which the intervention is perceived as likely to achieve its purpose” | The perceived benefits of injections | “Just keeping my eyesight is the main benefit, and watch[ing] the grandchildren grow up.” (P9)  “Well if it slows it down, that’s the main advantage if it slows down what is going on with my eye. That is to me the main thing. Hopefully then I would be able to see better than if I don’t have treatment. If I didn’t have the injections, it may go a lot quicker. That’s the one thing I can think of off the top of my head.” (P14)  "She [the participant, the speaker’s mother] just wants to live the rest of her days and moderately see. So anything that can do that. Let’s say 5 years span. If it’s going to slow down the process, give her better quality of life, better vision of her left eye as well with the treatment, she will have it… She might go blind in a future but every little bit helps. So give her two to three years of vision so she can watch TV, read books. After that there is no miracle. After that she can live with it.” (P25) |
|  | Uncertainties about the effectiveness of treatment | “I saw the benefits of having the [wet AMD] injections, but I am not sure if I will get the benefit of this new one.” (P24)  “Well, I wouldn’t like to do it [*have the injections*] for nothing, to get no results at all. But I wouldn’t know until it’s done.” (P30) |
| Self-efficacy: “The participant’s confidence that they can perform the behaviour required to participate in the intervention” i.e. in the case of our project, feeling confident to attend injection appointments | Patients’ confidence in commitment to long-term treatment | [Relative speaking on behalf of participant] “Maybe if she did have a treatment and it wasn’t getting better or for some reason it was getting worse, she would think that it wasn’t getting better. She would probably say “I am going there every month and I still can’t see. So what’s the point of going?” But she wouldn’t think that over the first 6 months or so. I would say, give it some time. Because even when she was having injections in her right eye she was like, “I can’t see” and every time she was having injection it would become clearer. Not too much, a little clearer. And then she would get her confidence back.” (P25) |
|  | Preference for injections in hospital vs in community [and how these impact self-efficacy] | “I think my optician is very good, he has been very helpful. But I think I would still feel more content with coming to hospital. I think there is more people on hand if anything goes wrong, at the opticians, it’s only him. So at hospital it will give me more confidence.” (P10) |
| ***Non-TFA categories and codes*** | | |
| Inter-participant differences | Differences linked to clinical profile/GA stage | *“I've had this [GA] since the mid-'90s. So I suspect I'm at the end of the spectrum. If I were earlier on, I certainly would be going for the most aggressive treatment.”* (P26) |
|  | Differences linked to socio-demographic characteristics [age, participant’s living/family situation, socioeconomic status etc] | *“It must be difficult if you’ve got no one and you haven’t got enough money and you’re not very well off to get cabs, et cetera, it must be difficult for them, especially with impaired vision, to get to and from the hospital, getting on buses and trains, et cetera. That’s what I find is their disadvantage but it’s not me.”* (P13)  *“I chose no injection options because I am old enough. If I carry on for another four years, I don’t care what happens after that… For younger people it’s different. They are the ones to get old and blind. If they are young, for them it’s better than me. I am old enough. If I lose my eye it’s too bad, but I have done my-- you know what I mean.”* (P22) |
|  | Differences linked to intravitreal injection history | *“It’s better to get something done. You see, because I had all those injections, so many in this eye, I know how it does feel and how it happens. That’s why I don’t mind having injections.”* (P2) |
|  | Relatives' and/or friends' eye history/treatment and their influence on participants  [This is about how knowing other people’s experiences of injections/treatment etc affects the participant] | *“My neighbour is having injections and he is very happy too. I ask questions about people, it makes me more comfortable to find out about other people.*” (P8)  “*I myself wouldn't want one [an injection]. But I would have one now, because I've seen my dad have them. I see that they don't hurt. I think it's only people that haven't had an injection that would be worried. Because he's used to them now, he's fine.*” (P16) |
| Impacts of GA on quality of life | Functional impacts | “*I can’t rely on my eye sight anymore. It’s restricted, I suppose, certain things for me. There is lots of different things that I can’t do now that I used to happily do. I used to do a lot of cooking and I am not doing it now. Like sometimes I put something in the saucepan and I am stirring it but I may not actually see what am I stirring or like potatoes coming to a boil. Things like that.*” (P14) |
|  | Emotional impacts | *“I do feel angry and depressed sometimes but I have no choice. I have to accept it.”* (P2) |
|  | Impacts on family and social life | *“[If I have treatment] my family will be happy that it’s not getting worse. They will be worried if I can’t see very well. If it gets worse they will help me with self-care, hospital appointments and so on.”* (P12) |
| Trust in professionals | Trust in professional judgement of the value of treatment | *“I don’t mind having injections. If doctors say it will help me, I don’t mind having injections.”* (P8)  *“There is nothing that would put me off. The doctors know better. If they think I need it I am happy to come for injection.”* (P5) |
| Longer-term hopes and wishes for treatment | Motivations/goals for treatment now | *“[I would hope] that I keep my sight until I die… Whatever sight it might be. My mum went completely blind so I just want to keep my sight as long as I can and that’s it.”* (P13) |
|  | Wishes and expectations for future treatments | *“If I could keep whatever [vision] I have that would be very excellent, if you can stop it there and it doesn’t get worse.*” (P28)  *“You know, in a year’s time, it [the available treatments] could be different again. It could be that we could get one injection every six months. It might not work out like that, but it rolls on so quickly these days.”* (P9) |
| Knowledge and understanding about GA | Knowledge and understanding about GA, as a potential rationale for more education | *“I think this has been really helpful, having this conversation. Because after reading that pack at home I got very upset, very stressed out, thinking that I am going blind. All of a sudden I turn to have a problem if what I was thinking was my good eye. So today has been very useful, making thing face-to-face it helped me to understand. And I think my mother having thrombosis and losing her eyesight and seeing all of that… I don’t want to go blind, I really don’t.”* (P10) |
| Thoughts about research and trial processes | Thoughts/reflections on our pilot project | *“There is lots of information you give out anyway and the explanations… It’s quite good enough.*” (P20) |
|  | Thoughts/reflections on GA research and trials more generally | “*I would have to consider the treatment. But when I was offered to participate in other trial, I didn’t know If I was going to have the treatment, I could be the one that wasn’t getting it. So I kind of thought it will be waste of my time.*” (P21) |
