## Supplementary figures and images for "Exploring patient acceptability of emerging intravitreal therapies for Geographic Atrophy: a mixed-methods study"

### Appendix 4 - OCT images for Participant 4

**Appendix 4: OCT images for Participant 4**

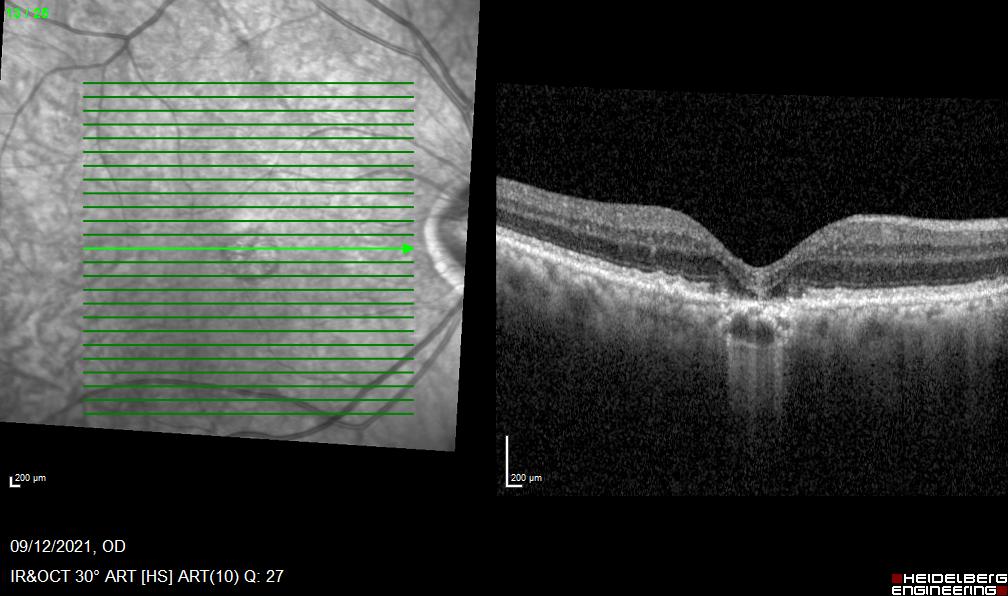
**Right eye**

**Left eye**

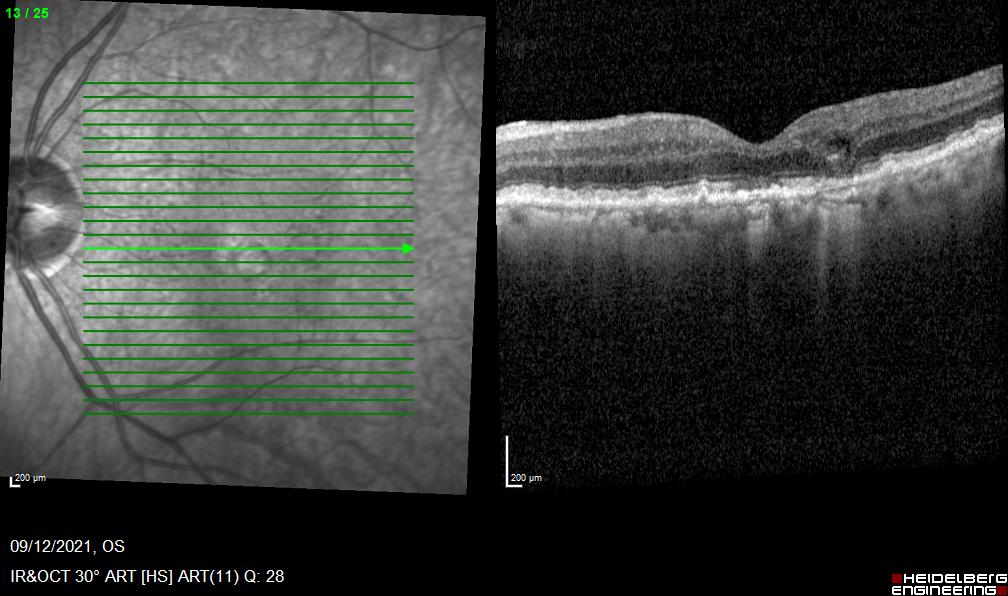
